# Segmentation of metabolically relevant adipose tissue compartments and ectopic fat deposits

**DOI:** 10.64898/2026.02.25.26347069

**Authors:** Tobias Haueise, Jürgen Machann

**Author notes:** <u>Contact:</u>.

## Abstract

Chemical shift-encoded magnetic resonance imaging using high-resolved 3D Dixon techniques enables the non-invasive and radiation-free assessment of whole-body adipose tissue and ectopic fat distribution. Automatic deep learning-based segmentation of metabolically relevant adipose tissue compartments and ectopic fat deposits in parenchymal tissue is the most important image processing step for the quantification of adipose tissue volumes and ectopic fat percentages from whole-body imaging.

This work presents a segmentation model dedicated to the segmentation of 19 metabolically relevant adipose tissue compartments and ectopic fat deposits from whole-body Dixon MRI. The trained segmentation model is available upon request. Related post-processing routines to compute volumes and fat percentages are publicly available: https://github.com/tobihaui/WholeBodyATQuantification.

## Introduction

During the last years, magnetic resonance imaging (MRI)-based volumetric quantification of adipose tissue has become an important tool for the estimation of potential risk of developing metabolic diseases. There is evidence that visceral adipose tissue (VAT) and ectopic lipid accumulation in parenchymal tissue of organs such as the liver or pancreas are particularly involved in the pathogenesis of type 2 diabetes.^1^ Dedicated MR examinations are performed in several large-scale cross-sectional and longitudinal studies (e.g., the UK Biobank^2^ or the German National Cohort [NAKO]^3^) and in various interdisciplinary research projects. The German Center for Diabetes Research (DZD) runs multiple clinical studies including MRI examinations for basal phenotyping of participants at increased risk for metabolic diseases and follow-up after (individualized) interventions.

This work was done with a special focus on bridging the gap between advanced MRI as, for example, assessed in single- or multi-centric studies within the DZD and the (clinical) need for standardized imaging-based outcomes. The nnU-Net framework, as proposed by Isensee et al.^4^, offers the possibility to train a strong baseline model focused on application without the need for manual tuning of model specifications.

The aim of the work was to develop a model based on a carefully curated training dataset that segments the adipose tissue compartments and ectopic fat deposits relevant in the context of clinical studies in diabetes research of the DZD and beyond.

## Materials and methods

### MRI acquisition

Two-point and multi-echo Dixon sequences are available on most clinical MR-units, independent on manufacturer or magnetic field strength. Standard two-point techniques enable generation of fat and water selective images by acquiring in-phase and opposed-phase images. This allows reliable differentiation of adipose and lean tissue and determination of fat fraction (FF) by calculating the ratio of fat and water within the respective region of interest. In contrast to this simple initial approach, multi-echo Dixon sequences acquire 6 (or more) echoes, hereby allowing for calculation of proton density fat fraction (PDFF) by incorporating the effective transverse relaxation time T2* and using a low flip angle to avoid T1-related bias. With this, the absolute percentage of fat and its distribution can be assessed in organs/tissues of interest within short measurement time during a single breath-hold.

Whole-body Dixon imaging is usually being performed in 6-7 slightly overlapping blocks with 80-100 partitions each (total measurement time including localizer imaging about 10 min) and these blocks are finally combined to a seamless whole-body dataset. For acquisition of the trunk, 2-3 blocks ranging from shoulders to thighs are sufficient. Study participants are lying in supine position on the patient table with mounted spine-array coil and the body is covered with two body-array-coils and a multi-channel peripheral angio-coil on the lower legs.

### Classes

Figure 1 illustrates all 19 classes included in the model. Pairwise structures, for example, kidneys or muscle groups and multi-instance classes, for example, the vertebral bodies, are classified in one class and will be separated during post-processing/quantification. Similarly, subcutaneous adipose tissue is one large class that can be separated into, for example, thoracic, abdominal, and gluteofemoral sub-compartments during post-processing/quantification for more detailed analysis. For simplicity, due to image resolution and in view of generalizability reasons, muscles are classified in functional groups, for example, Mm. gluteus maximus, gluteus minimus, gluteus medius, and piriformis are classified in one class *Glutes*.

**Figure 1:**
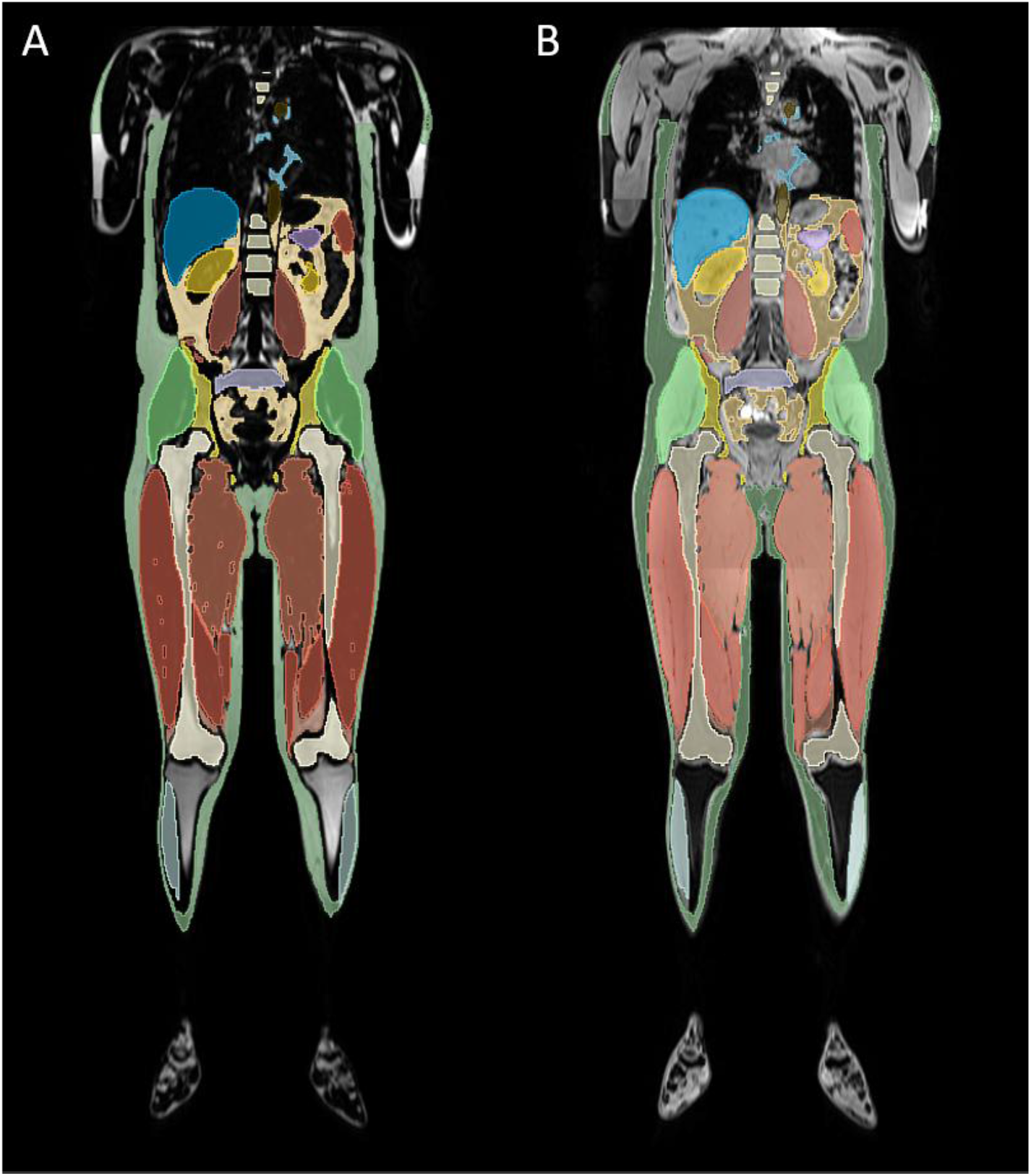
Overview of segmented classes on fat-(A) and water-selective (B) Dixon outputs. Class *Back muscles* is not visible

In parenchymal organs (liver, pancreas, bone marrow and skeletal muscle), PDFF as well as the area of macroscopic fat accumulation, i.e., intermuscular fat or inter-lobular fat in the pancreas, are determined. Additionally, volumes (or related structural measures, e.g., maximum cross-sections) of skeletal muscle and organ parenchyma can be measured.

### Model training

MR datasets from n = 76 self-identified White individuals (n = 48 women) between 20 and 78 (mean 46.1 +/-16.3) years and BMI ranging from 19.7 to 41.2 (mean 27.7 +/−5.1) kg/m^2^ were used to train five cross-validation folds of a 3D full-resolution residual encoder (*ResEnc M* configuration^5^) U-Net model for the segmentation of 19 adipose tissue compartments and ectopic fat deposits for 1000 epochs within the nnU-Net framework.^4^ The ground truth was curated based on outputs from previously developed models^6–9^ and computer-supported manual annotation.^10^ All ground truth segmentation masks have undergone rigorous visual inspection and manual correction/optimization for all classes.

Evaluation of the training process in terms of mean and standard deviation over all five folds of cross-validation involves a combination of overlap-based (Dice Similarity Coefficient [DSC], Intersection over Union [IoU]), boundary-based (Normalized Surface Distance [NSD]) and application-specific (Mean absolute volume error) metrics.^11^

All MRI examinations have been approved by the ethics committee of the University of Tübingen. All participants gave written informed consent. All data have been pseudonymized.

## Results

In model training, fast convergence was obtained for all cross-validation folds. Mean pseudo DSC during all epochs training over all cross-validation folds is shown for all classes in Figure 1. Figure 2 highlights the training process in the first 25 epochs. Cross-validation metrics for all classes are summarized in Table 1. DSC and IoU ranged from 0.872 +/-0.03 / 0.774 +/-0.04 for the *Sacrum* class to 0.989 +/-0.01 / 0.979 +/-0.01 for the *Subcutaneous Adipose Tissue* class. Volumetric error was below 3% for 14 out of 19 classes and below 5% for all but one class (*Pancreas*, 9.1 +/-11.1%). Bland-Altman analysis found low bias (mean / 1.96 SD < 0.05) in predicted volumes compared to ground truth volumes for 11 classes. From eight classes with moderate bias, six classes (*Visceral Adipose Tissue, Thigh Extensors, Iliopsoas, Aorta, Kidneys, Calves*) got overestimated, the other two classes (*Back muscles, Pancreas*) got underestimated by the segmentation model.

**Table 1:**
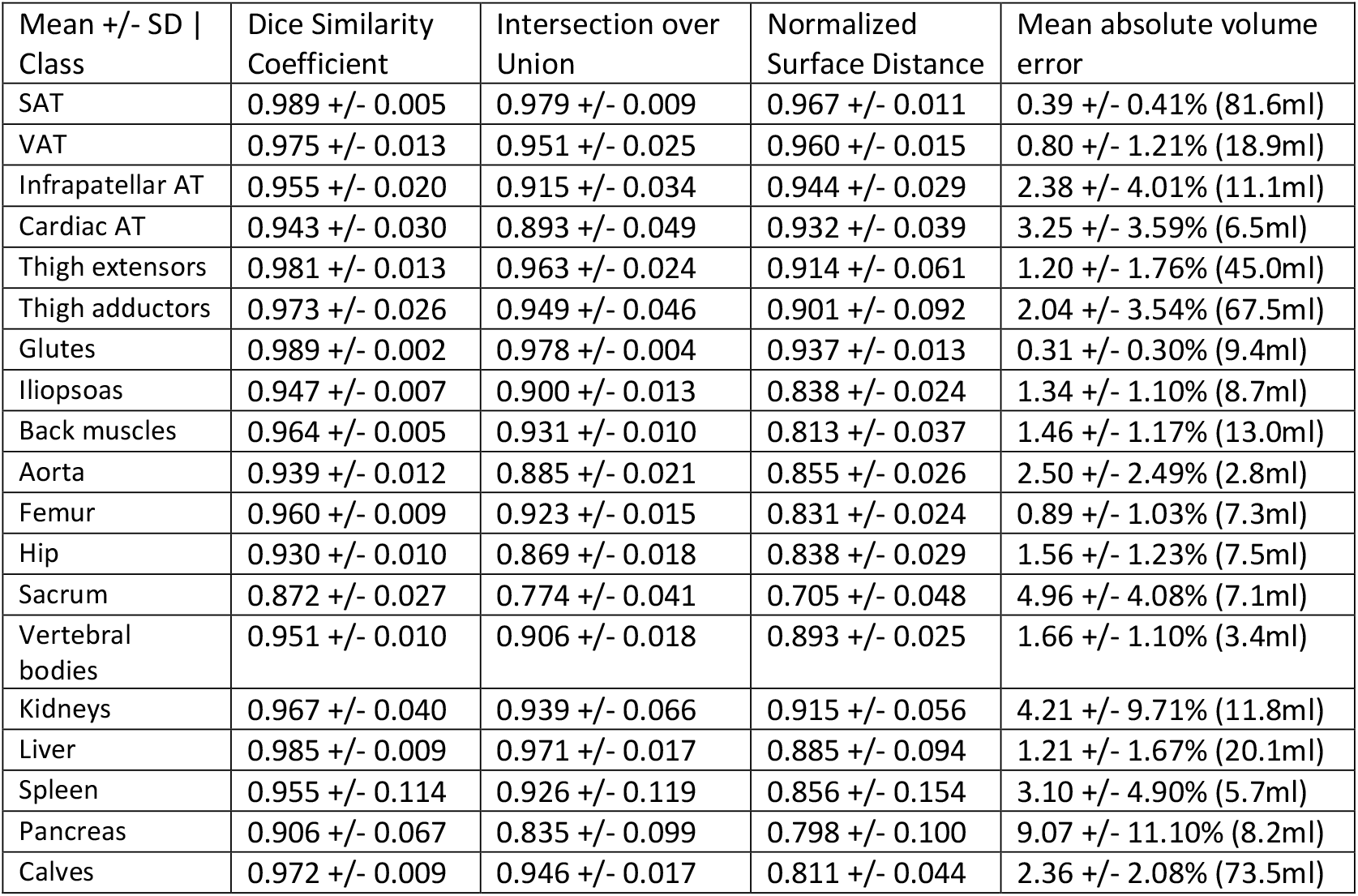
Five-fold cross-validation metrics presented as mean +/−standard deviation. Validation folds included 15 to 16 datasets.

| Mean +/- SD \| Class | Dice Similarity Coefficient | Intersection over Union | Normalized Surface Distance | Mean absolute volume error |
| --- | --- | --- | --- | --- |
| SAT | 0.989 +/- 0.005 | 0.979 +/- 0.009 | 0.967 +/- 0.011 | 0.39 +/- 0.41% (81.6ml) |
| VAT | 0.975 +/- 0.013 | 0.951 +/- 0.025 | 0.960 +/- 0.015 | 0.80 +/- 1.21% (18.9ml) |
| Infrapatellar AT | 0.955 +/- 0.020 | 0.915 +/- 0.034 | 0.944 +/- 0.029 | 2.38 +/- 4.01% (11.1ml) |
| Cardiac AT | 0.943 +/- 0.030 | 0.893 +/- 0.049 | 0.932 +/- 0.039 | 3.25 +/- 3.59% (6.5ml) |
| Thigh extensors | 0.981 +/- 0.013 | 0.963 +/- 0.024 | 0.914 +/- 0.061 | 1.20 +/- 1.76% (45.0ml) |
| Thigh adductors | 0.973 +/- 0.026 | 0.949 +/- 0.046 | 0.901 +/- 0.092 | 2.04 +/- 3.54% (67.5ml) |
| Glutes | 0.989 +/- 0.002 | 0.978 +/- 0.004 | 0.937 +/- 0.013 | 0.31 +/- 0.30% (9.4ml) |
| Iliopsoas | 0.947 +/- 0.007 | 0.900 +/- 0.013 | 0.838 +/- 0.024 | 1.34 +/- 1.10% (8.7ml) |
| Back muscles | 0.964 +/- 0.005 | 0.931 +/- 0.010 | 0.813 +/- 0.037 | 1.46 +/- 1.17% (13.0ml) |
| Aorta | 0.939 +/- 0.012 | 0.885 +/- 0.021 | 0.855 +/- 0.026 | 2.50 +/- 2.49% (2.8ml) |
| Femur | 0.960 +/- 0.009 | 0.923 +/- 0.015 | 0.831 +/- 0.024 | 0.89 +/- 1.03% (7.3ml) |
| Hip | 0.930 +/- 0.010 | 0.869 +/- 0.018 | 0.838 +/- 0.029 | 1.56 +/- 1.23% (7.5ml) |
| Sacrum | 0.872 +/- 0.027 | 0.774 +/- 0.041 | 0.705 +/- 0.048 | 4.96 +/- 4.08% (7.1ml) |
| Vertebral bodies | 0.951 +/- 0.010 | 0.906 +/- 0.018 | 0.893 +/- 0.025 | 1.66 +/- 1.10% (3.4ml) |
| Kidneys | 0.967 +/- 0.040 | 0.939 +/- 0.066 | 0.915 +/- 0.056 | 4.21 +/- 9.71% (11.8ml) |
| Liver | 0.985 +/- 0.009 | 0.971 +/- 0.017 | 0.885 +/- 0.094 | 1.21 +/- 1.67% (20.1ml) |
| Spleen | 0.955 +/- 0.114 | 0.926 +/- 0.119 | 0.856 +/- 0.154 | 3.10 +/- 4.90% (5.7ml) |
| Pancreas | 0.906 +/- 0.067 | 0.835 +/- 0.099 | 0.798 +/- 0.100 | 9.07 +/- 11.10% (8.2ml) |
| Calves | 0.972 +/- 0.009 | 0.946 +/- 0.017 | 0.811 +/- 0.044 | 2.36 +/- 2.08% (73.5ml) |

**Figure 2:**
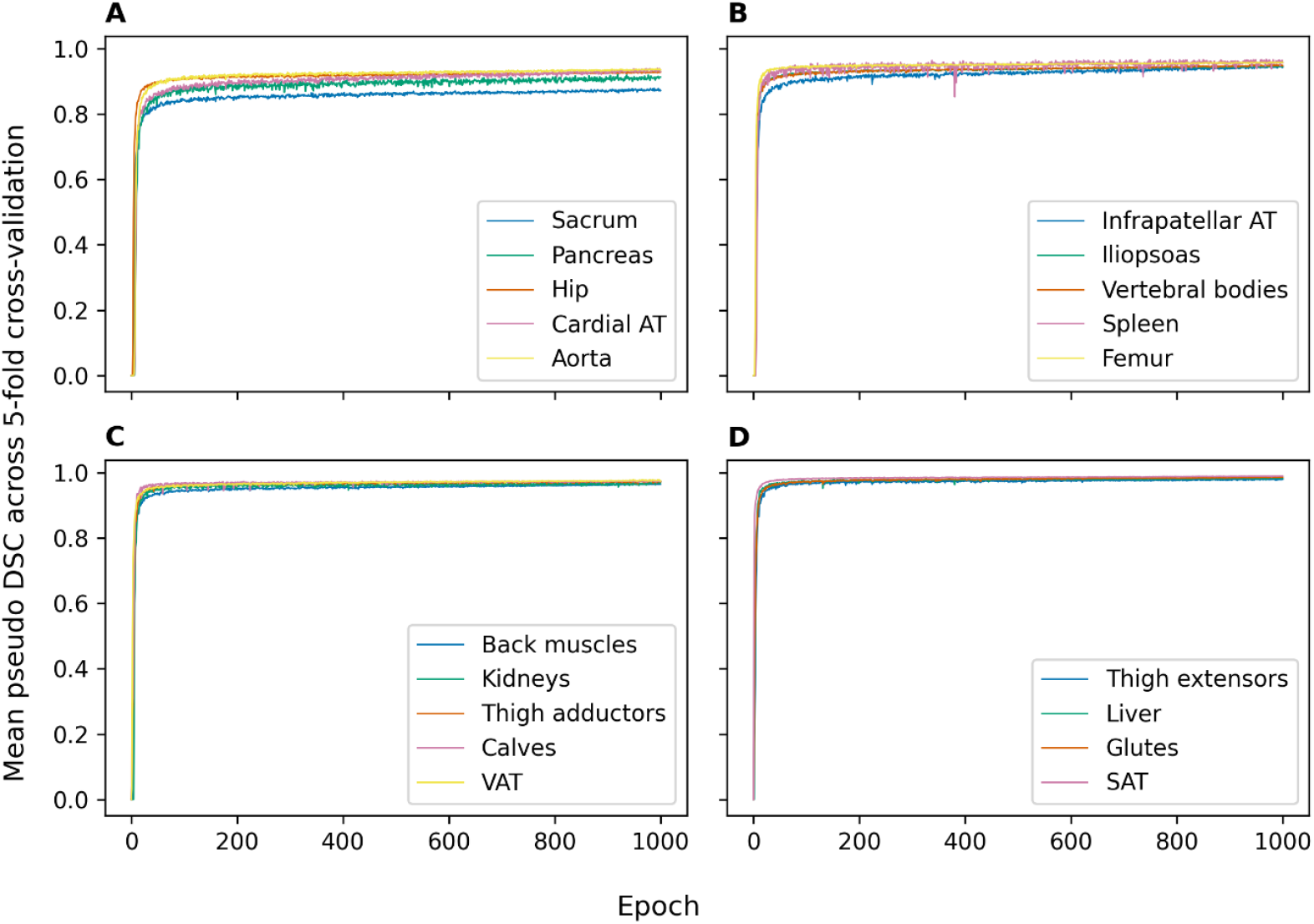
Mean pseudo Dice Similarity Coefficient during 1000 epochs of model training. Classes are arranged from lowest DSC (A) to highest DSC (D).

**Figure 3:**
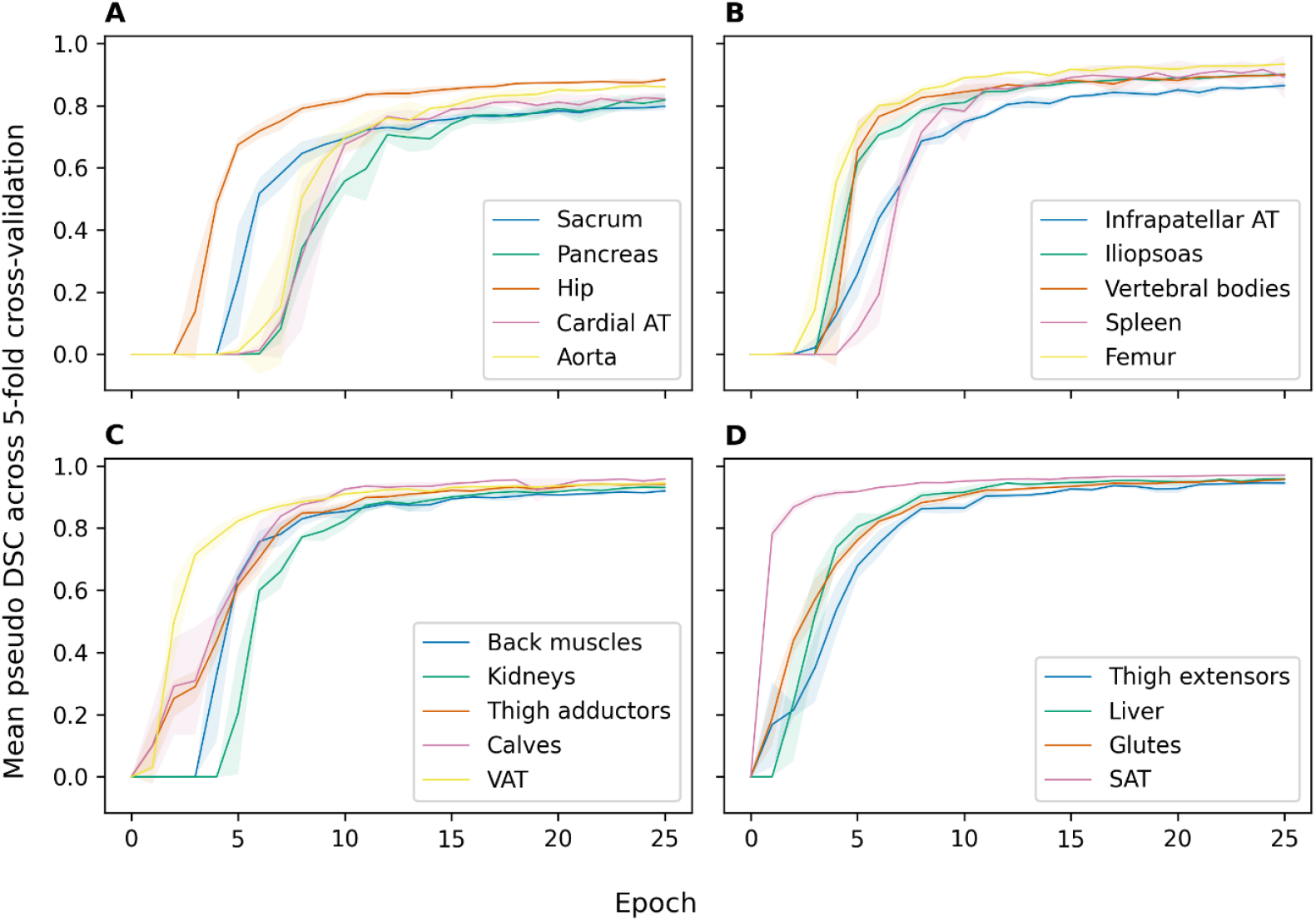
Mean pseudo Dice Similarity Coefficient during the first 25 epochs of model training. Classes are arranged from lowest DSC (A) to highest DSC (D).

Experiments were conducted using a private infrastructure, which has a carbon efficiency of 0.35 kgCO_2_eq/kWh (estimated data for 2024 from Umweltbundesamt for Germany). A cumulative of 210 hours of computation was performed on hardware of type A100 PCIe 40/80GB (Thermal design power of 250W). Total emissions are estimated to be 18.38 kgCO_2_eq of which 0% were directly offset. Estimations were conducted using the MachineLearning Impact calculator.^12^

## Discussion

This work presents a segmentation model that provides the masks for volumetric quantification of metabolically relevant adipose tissue compartments, ectopic fat deposits and skeletal muscle and organ volumes from whole-body Dixon MRI.

Given the increasing number of large “all-purpose” segmentation models available addressing a large number of anatomical target structures^9,13–17^, this work presents a small, application-specific alternative. One main advantage is the focus on metabolically relevant segmentation classes with carefully annotated ground truth labels, in contrast to coarse class definitions that, for example, “extended the visceral fat to match other organ borders.”^13^ Although being trained with a lower number of training datasets, cross-validation DSC of our model is comparable with the overview table provided in study by Graf et al.^13^ Especially in the large adipose tissue compartments, our model reached higher DSC (0.98 for both subcutaneous and visceral adipose tissue compared to 0.95 and 0.89 by Graf et al.^13^, respectively).

Further, the model only includes classes that directly result in MR-derived biomarkers for downstream analyses. These biomarkers directly contribute to real-world studies in diabetes research and complement standard clinical measures.^18–20^ The postprocessing routines to extract these biomarkers from the segmentation masks are included in the accompanying repository (https://github.com/tobihaui/WholeBodyATQuantification).

### Quantification of peri-X adipose tissue

Besides the basic postprocessing routines, more advanced methods are also included. For example, taking a segmented organ/anatomical structure as a starting point, the local amount of adipose tissue around that organ/structure can be evaluated using parametrizable dilation operations to focus on the organ’s/structure’s three-dimensional surrounding. This results in additional, anatomically-guided local adipose tissue volumes that we named *peri-X adipose tissue*, for example, peri-renal, peri-pancreatic, or peri-vascular adipose tissue.

### Limitations

The model has been trained using data from White individuals from southern Germany. While having paid close attention to include, besides sex, a broad age and BMI range to increase representativeness of the training dataset, an application of the model to another population should be carefully monitored. Further, the training dataset only included individuals without pathologically deformed organs.

## Conclusion

This work provides a carefully curated, application-specific segmentation model of metabolically relevant adipose tissue compartments, ectopic fat deposits, skeletal muscles, and organs, particularly useful in clinical studies including Dixon MRI.

## Data Availability

All data produced in the present study are available upon reasonable request to the authors. All code is available online at https://github.com/tobihaui/WholeBodyATQuantification.

## Appendix

**Appendix 1:**
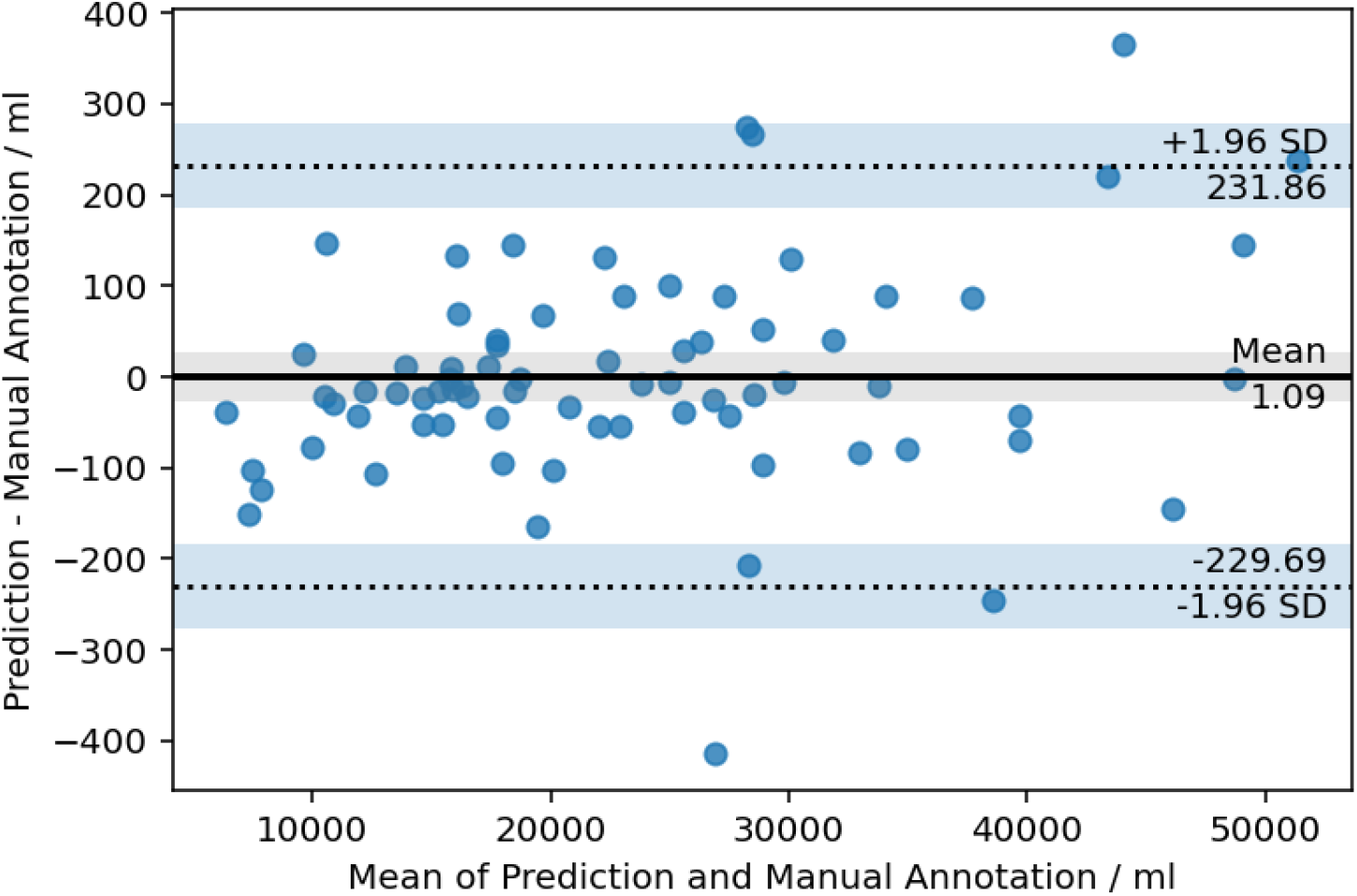
Bland-Altman plot for *Subcutaneous Adipose Tissue*

**Appendix 2:**
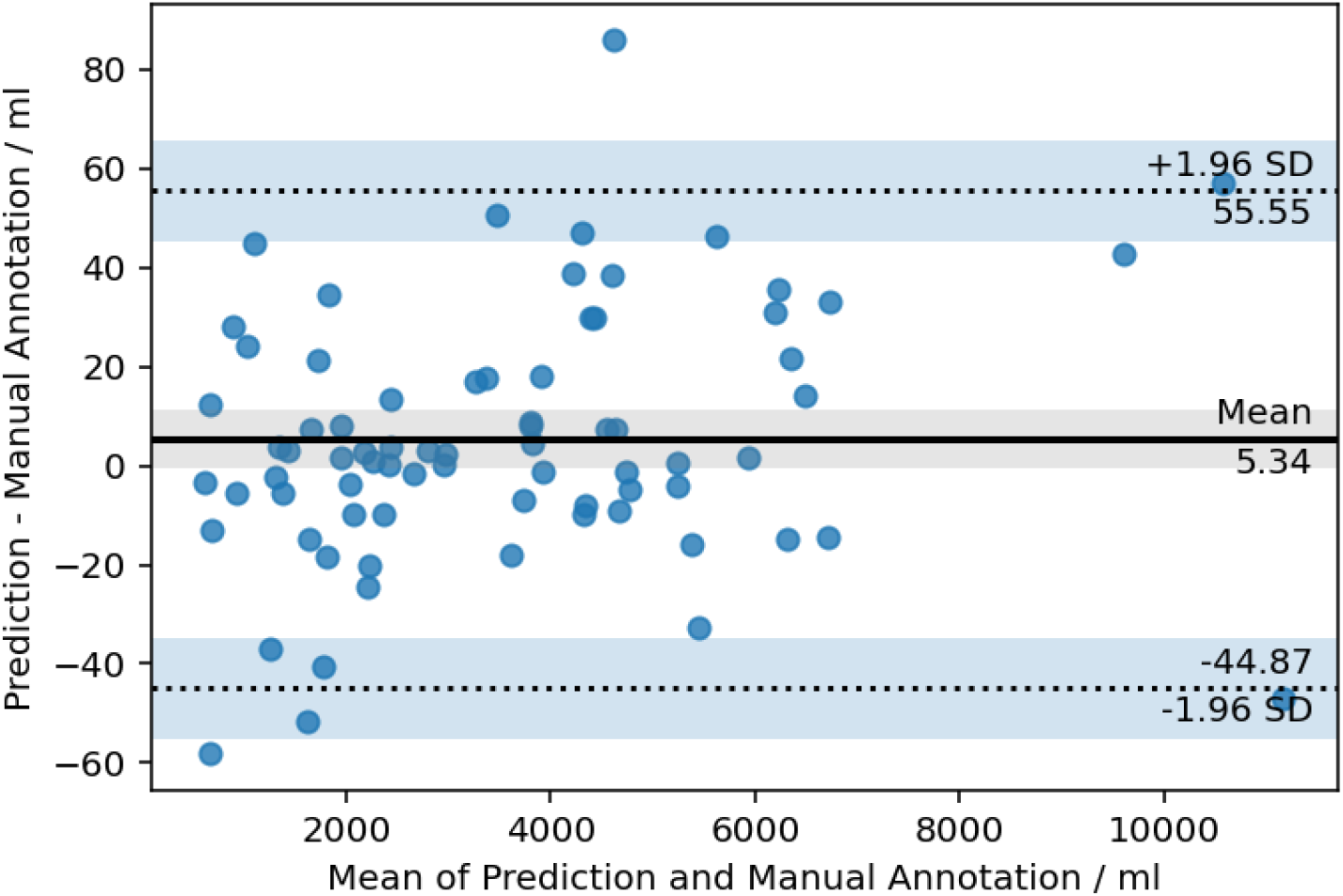
Bland-Altman plot for *Visceral Adipose Tissue*

**Appendix 3:**
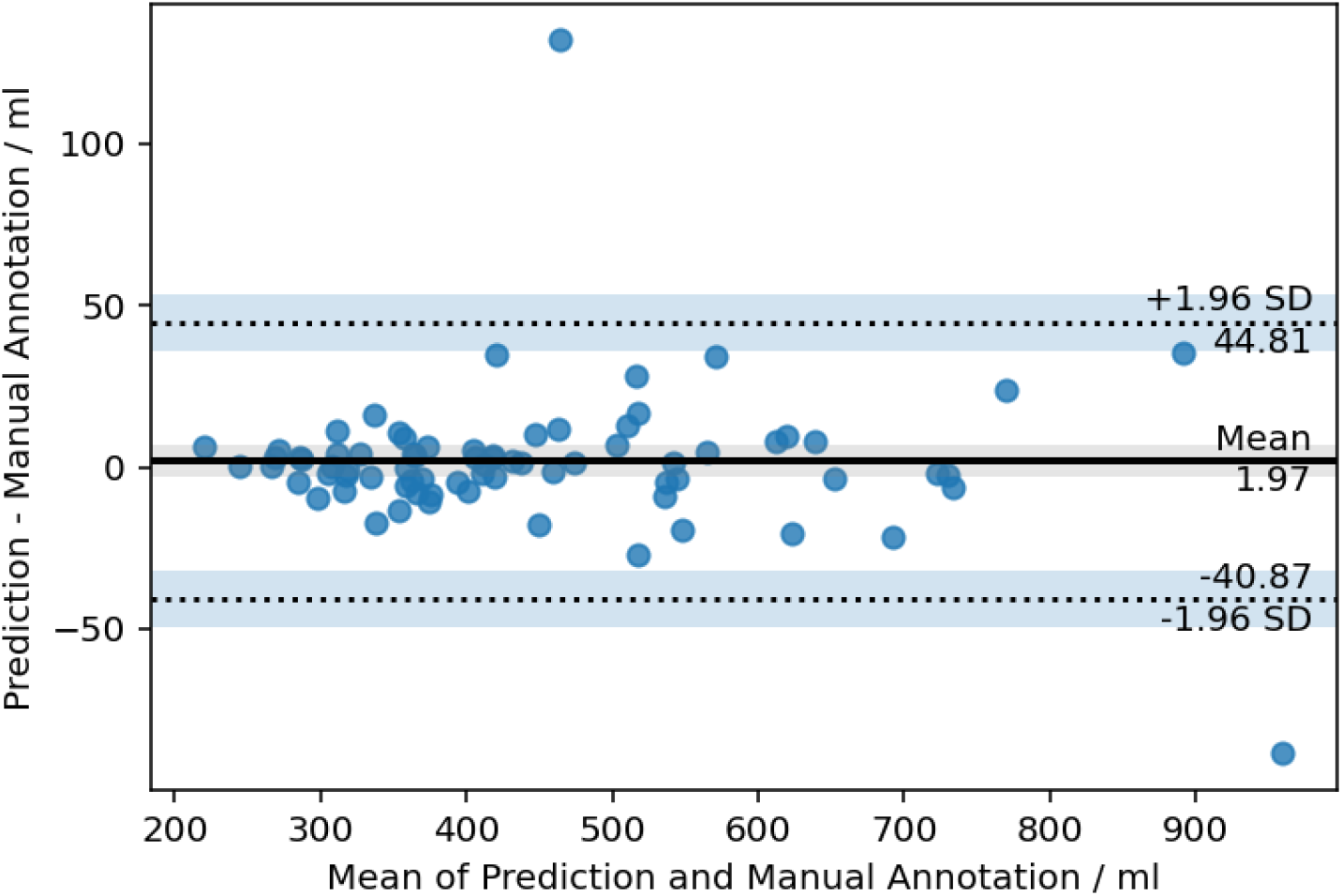
Bland-Altman plot for *Infrapatellar Adipose Tissue*

**Appendix 4:**
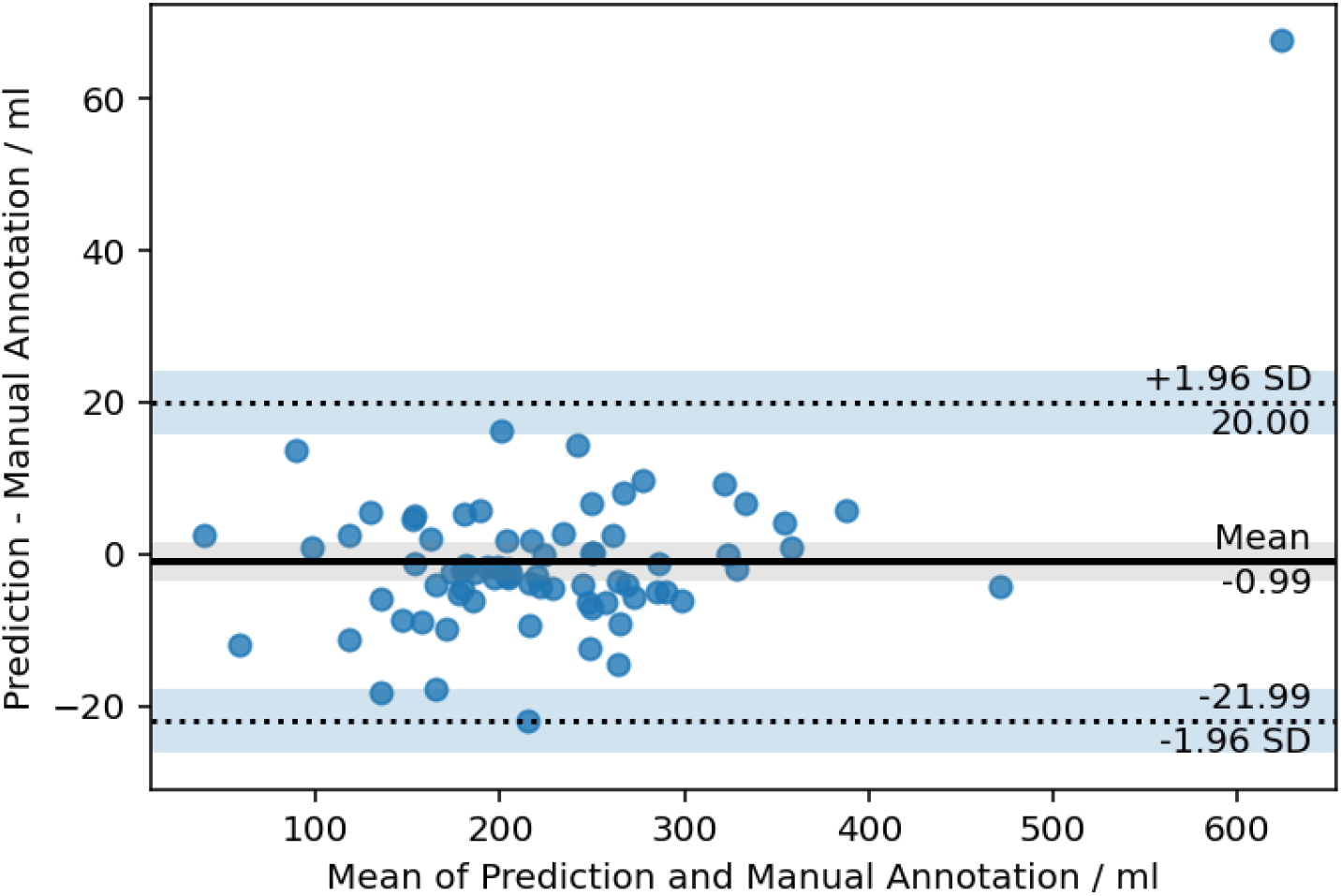
Bland-Altman plot for *Cardiac Adipose Tissue*

**Appendix 5:**
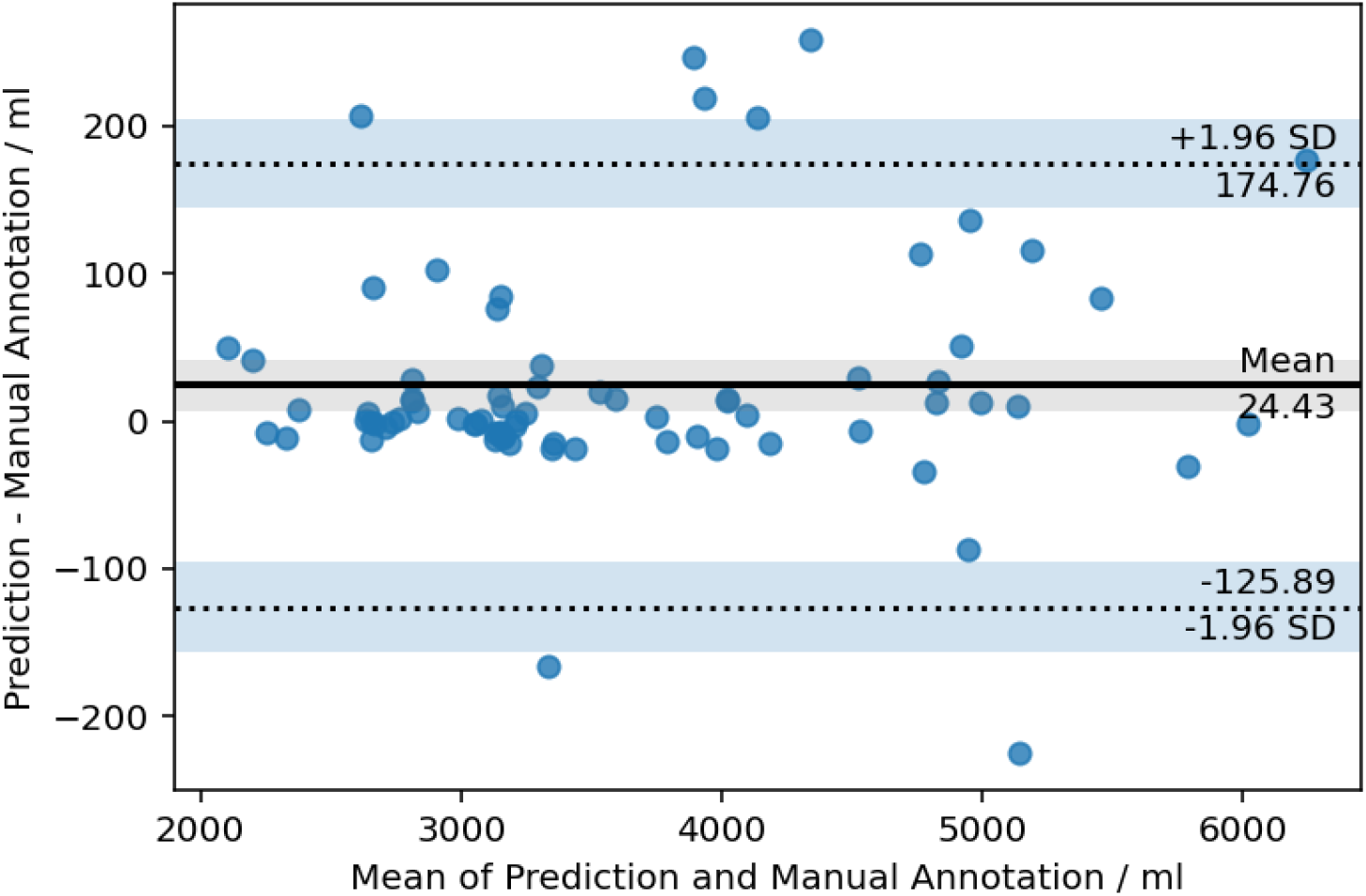
Bland-Altman plot for *Thigh Extensors*

**Appendix 6:**
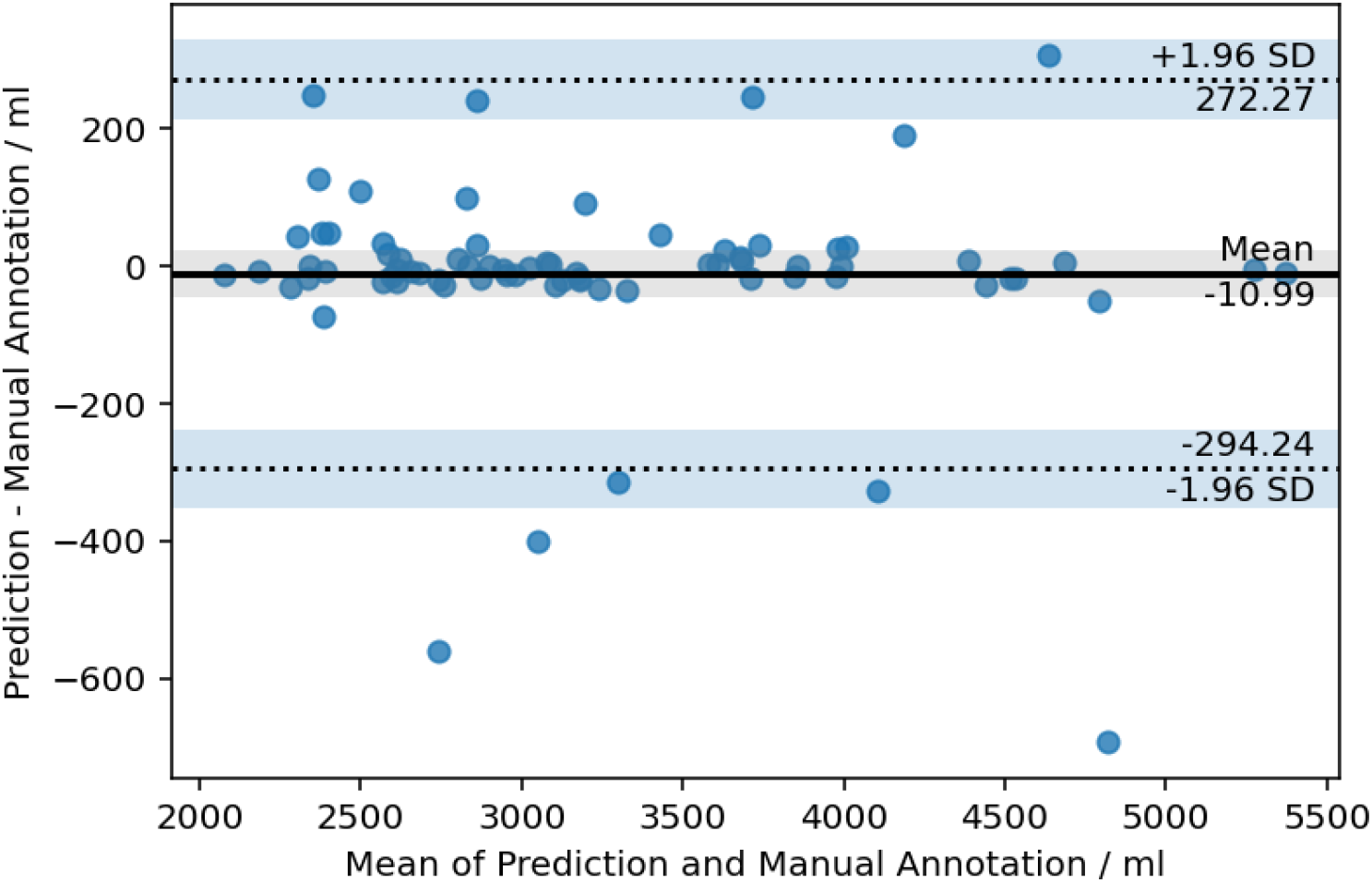
Bland-Altman plot for *Thigh Adductors*

**Appendix 7:**
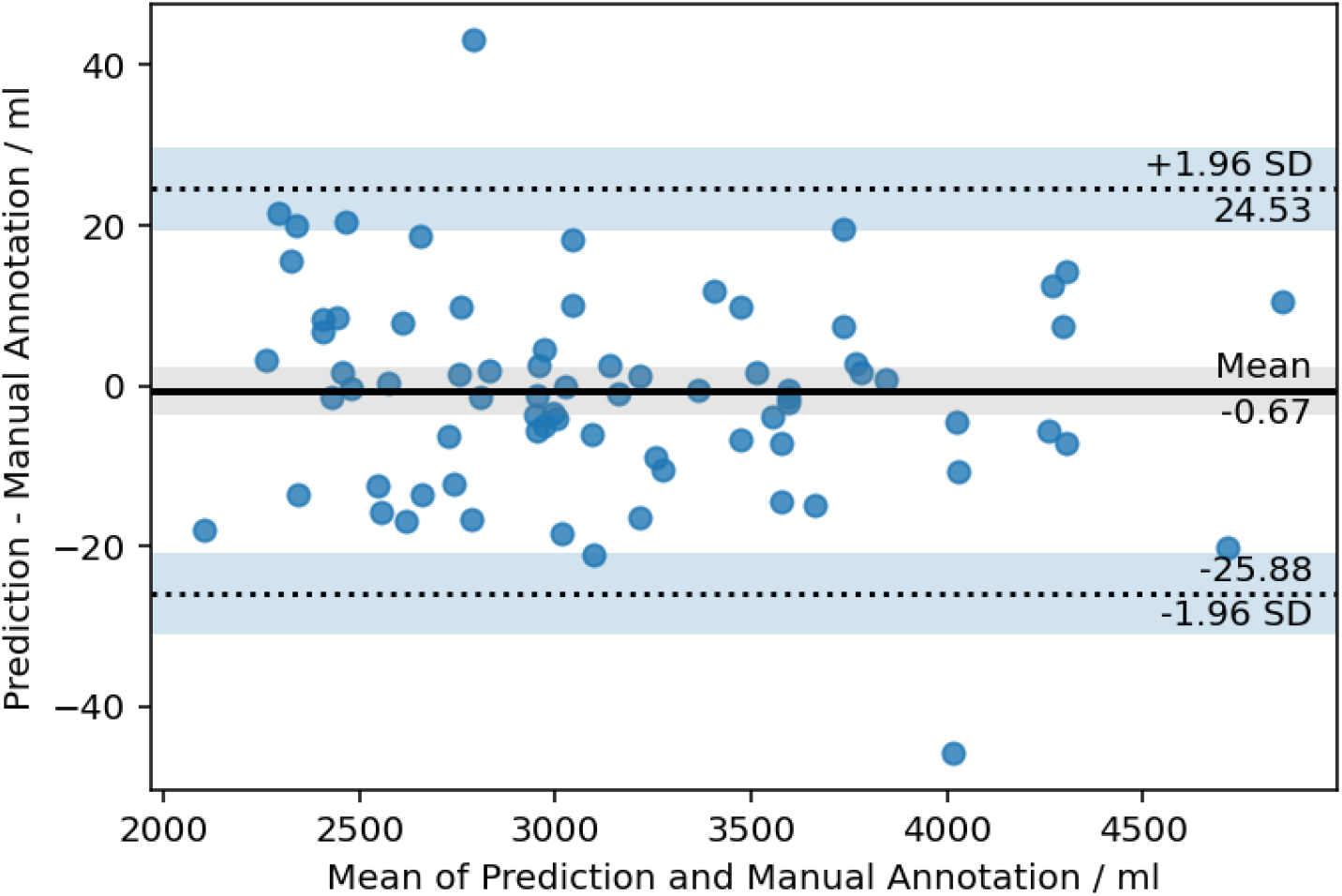
Bland-Altman plot for *Glutes*

**Appendix 8:**
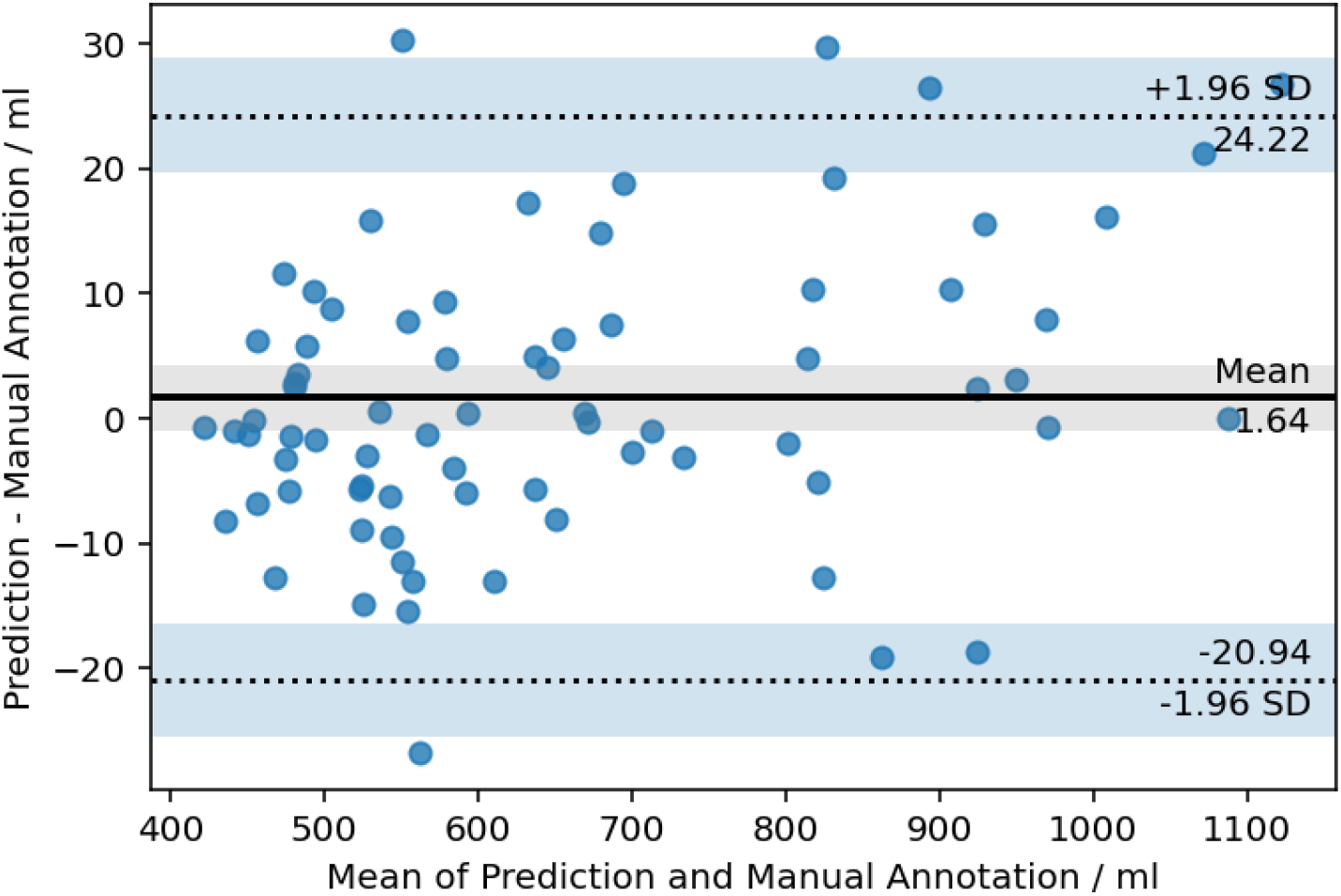
Bland-Altman plot for *Iliopsoas*

**Appendix 9:**
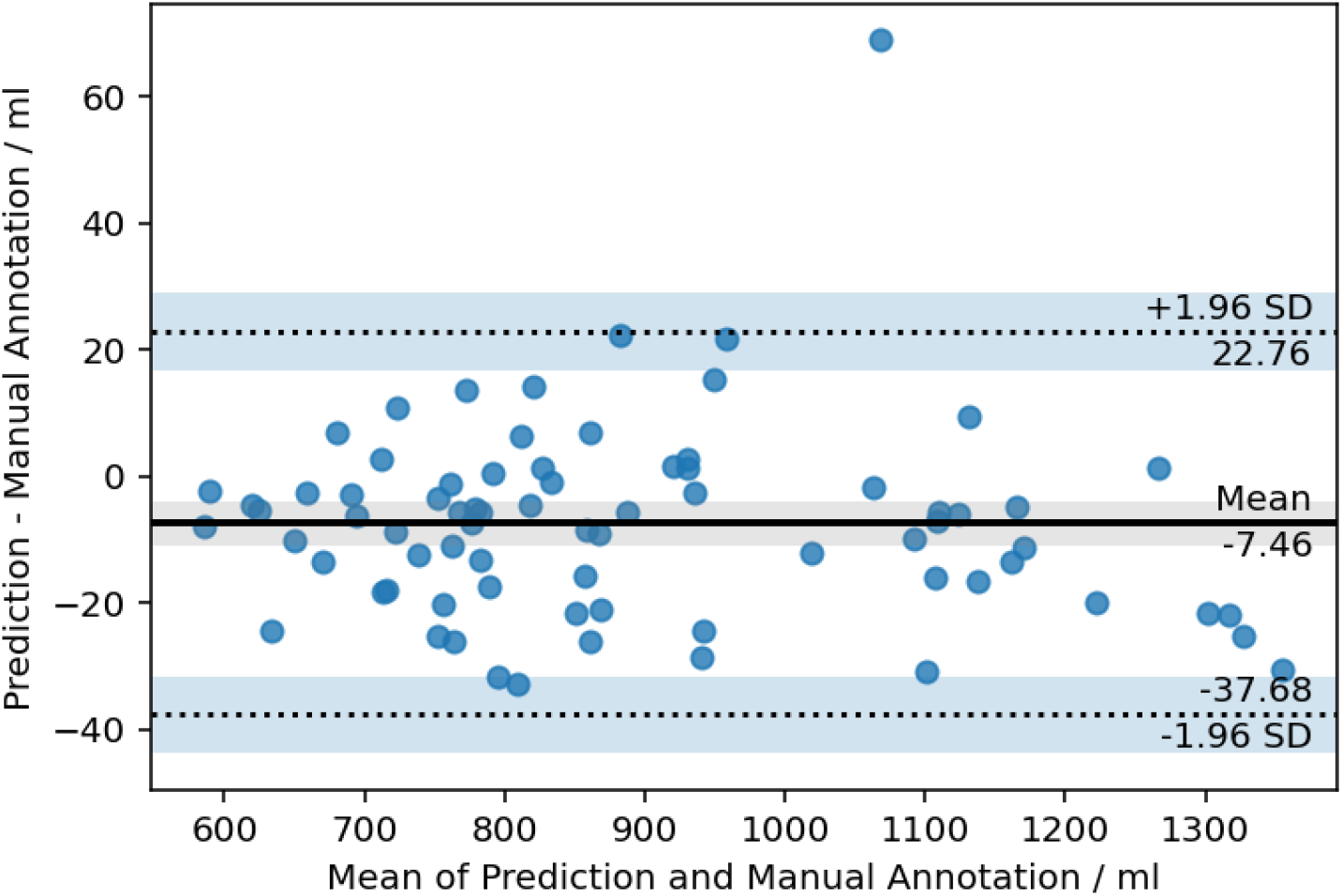
Bland-Altman plot for *Back muscles*

**Appendix 10:**
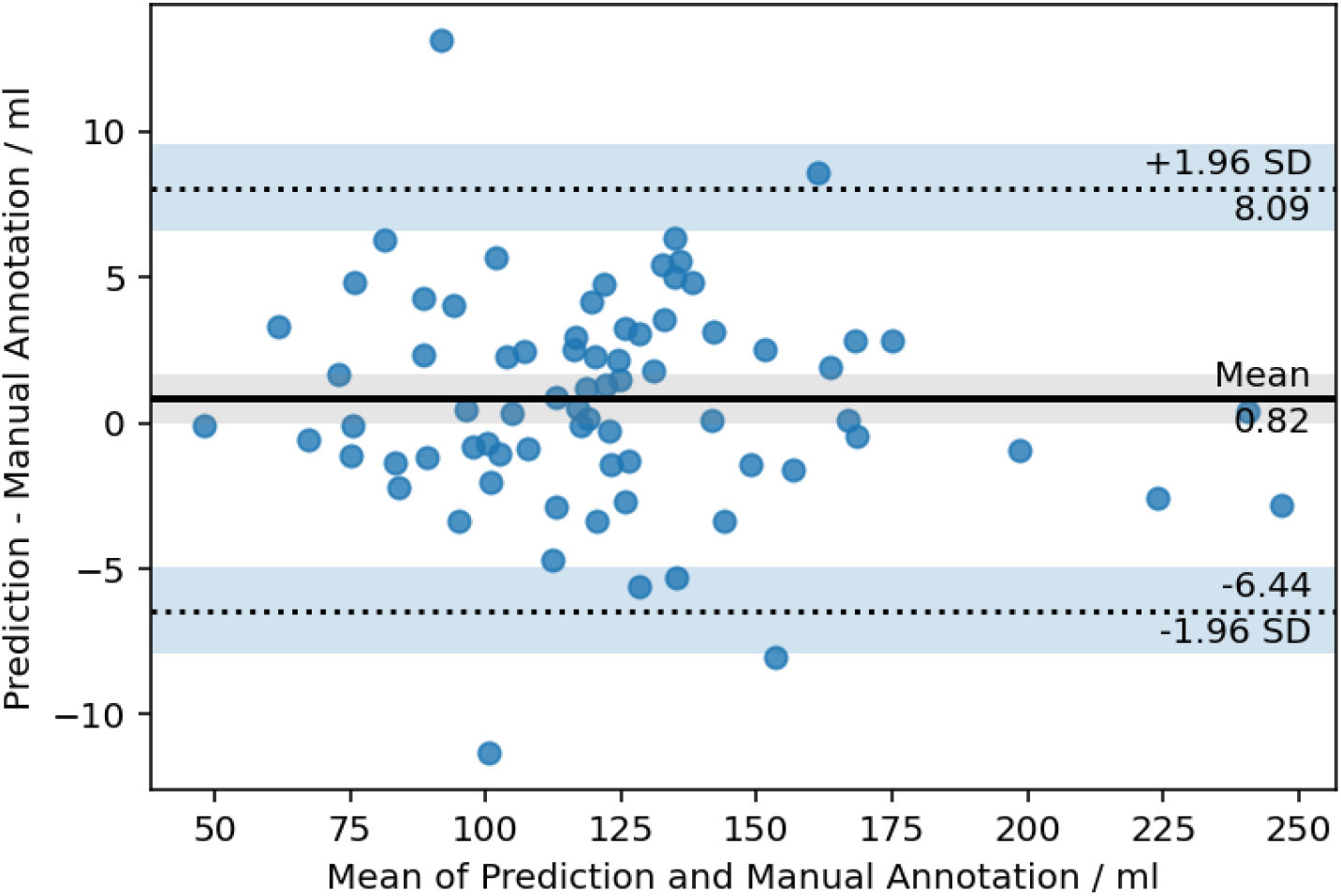
Bland-Altman plot for *Aorta*

**Appendix 11:**
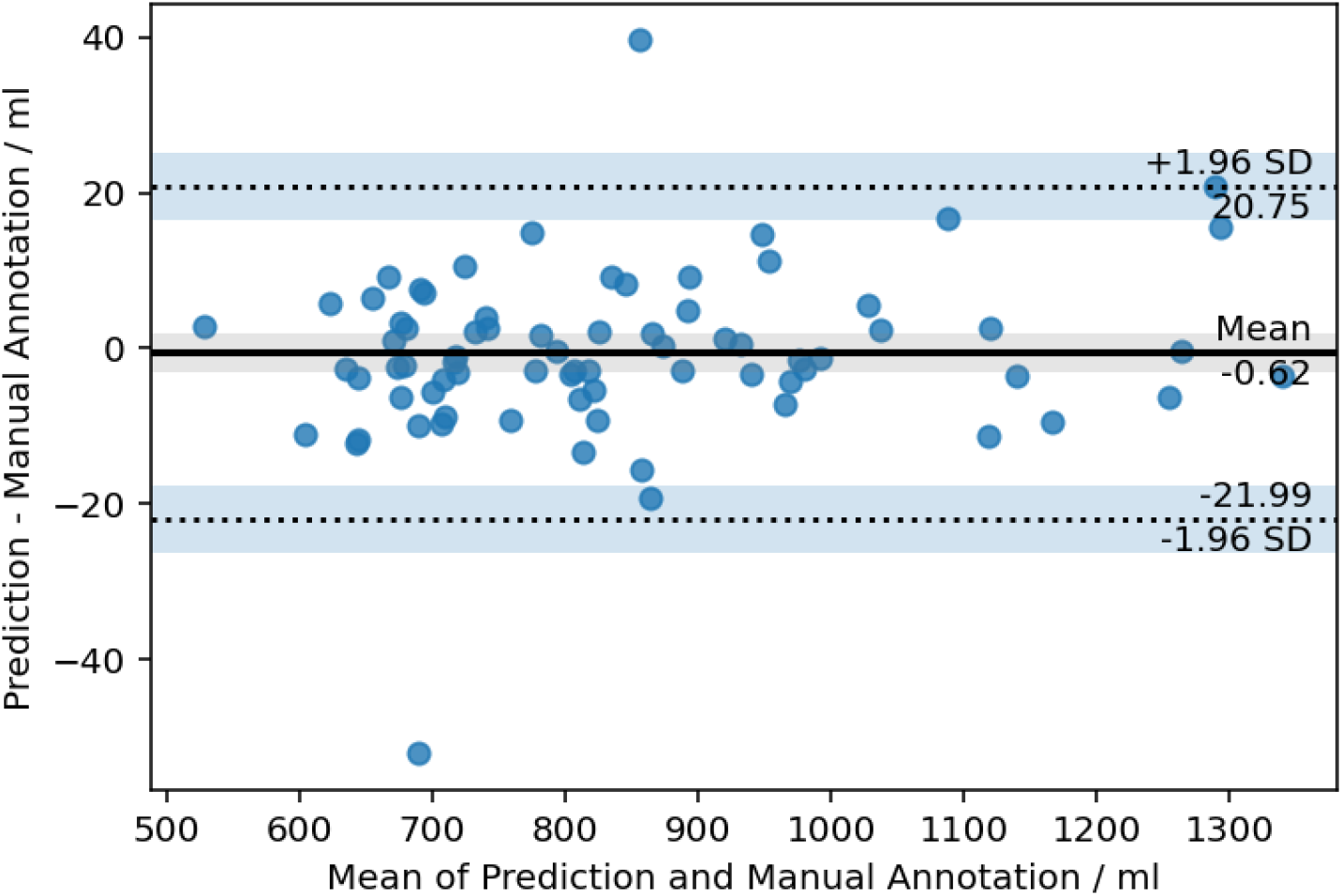
Bland-Altman plot for *Femur*

**Appendix 12:**
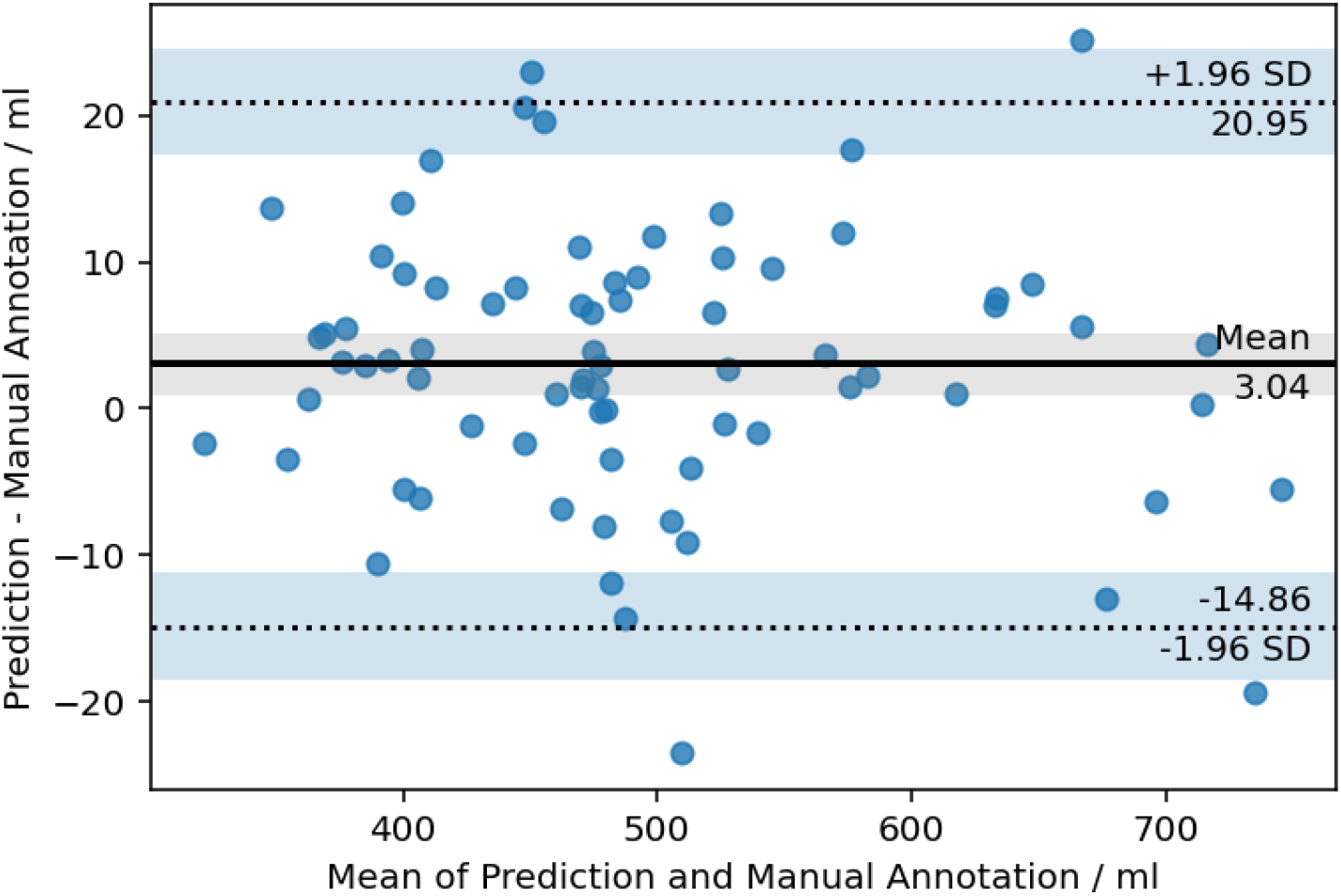
Bland-Altman plot for *Hip*

**Appendix 13:**
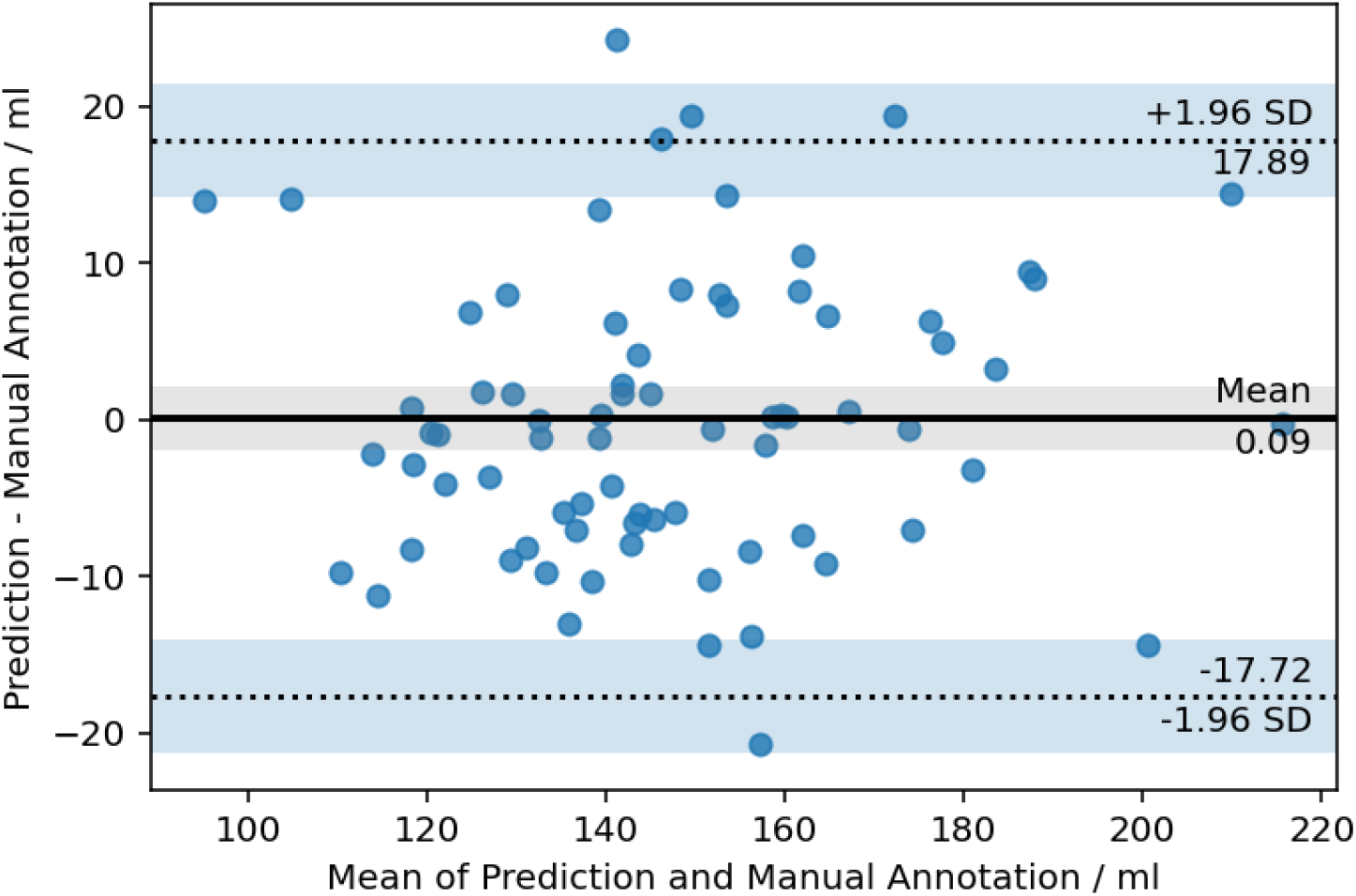
Bland-Altman plot for *Sacrum*

**Appendix 14:**
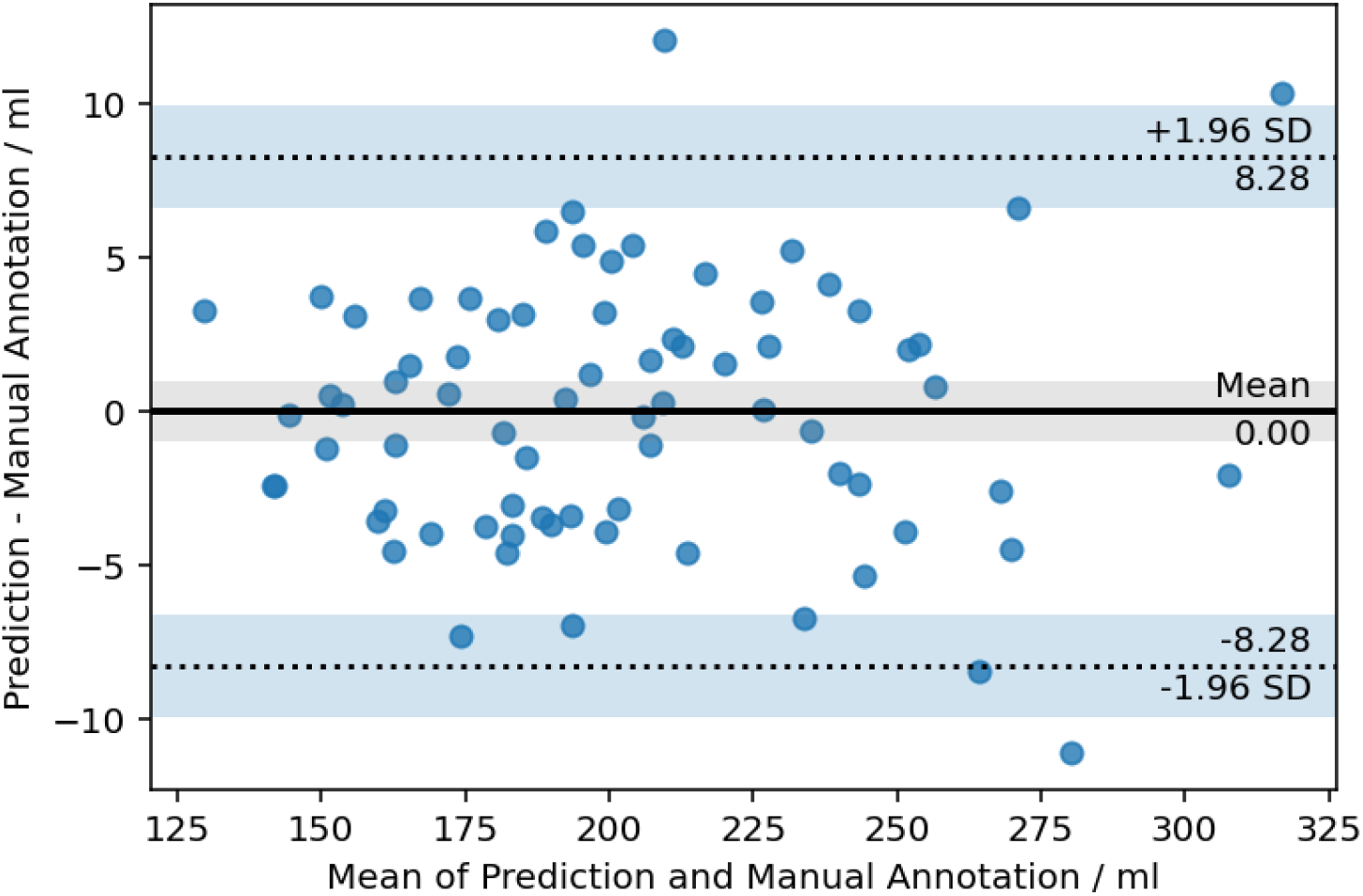
Bland-Altman plot for *Vertebral Bodies*

**Appendix 15:**
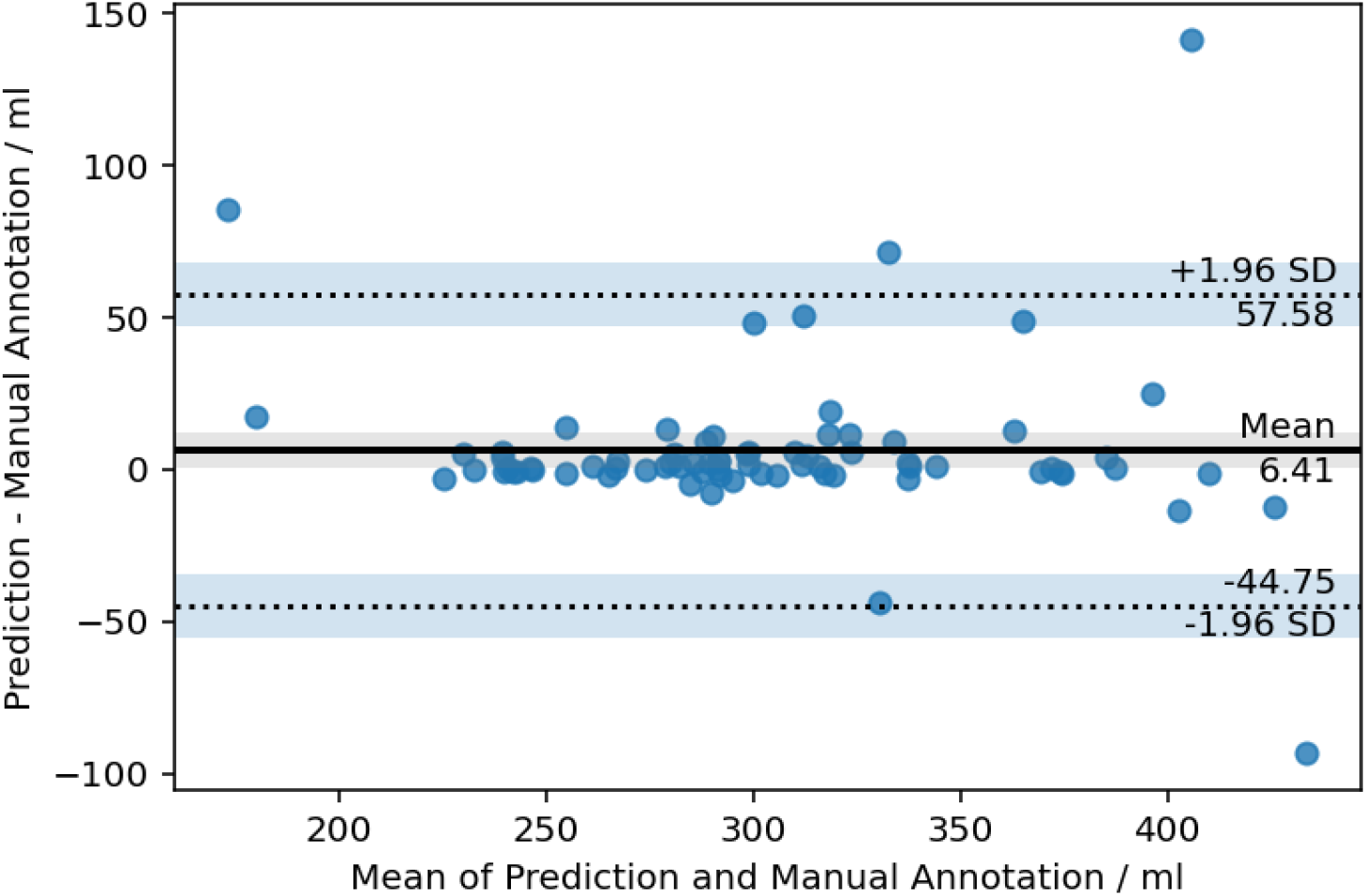
Bland-Altman plot for *Kidneys*

**Appendix 16:**
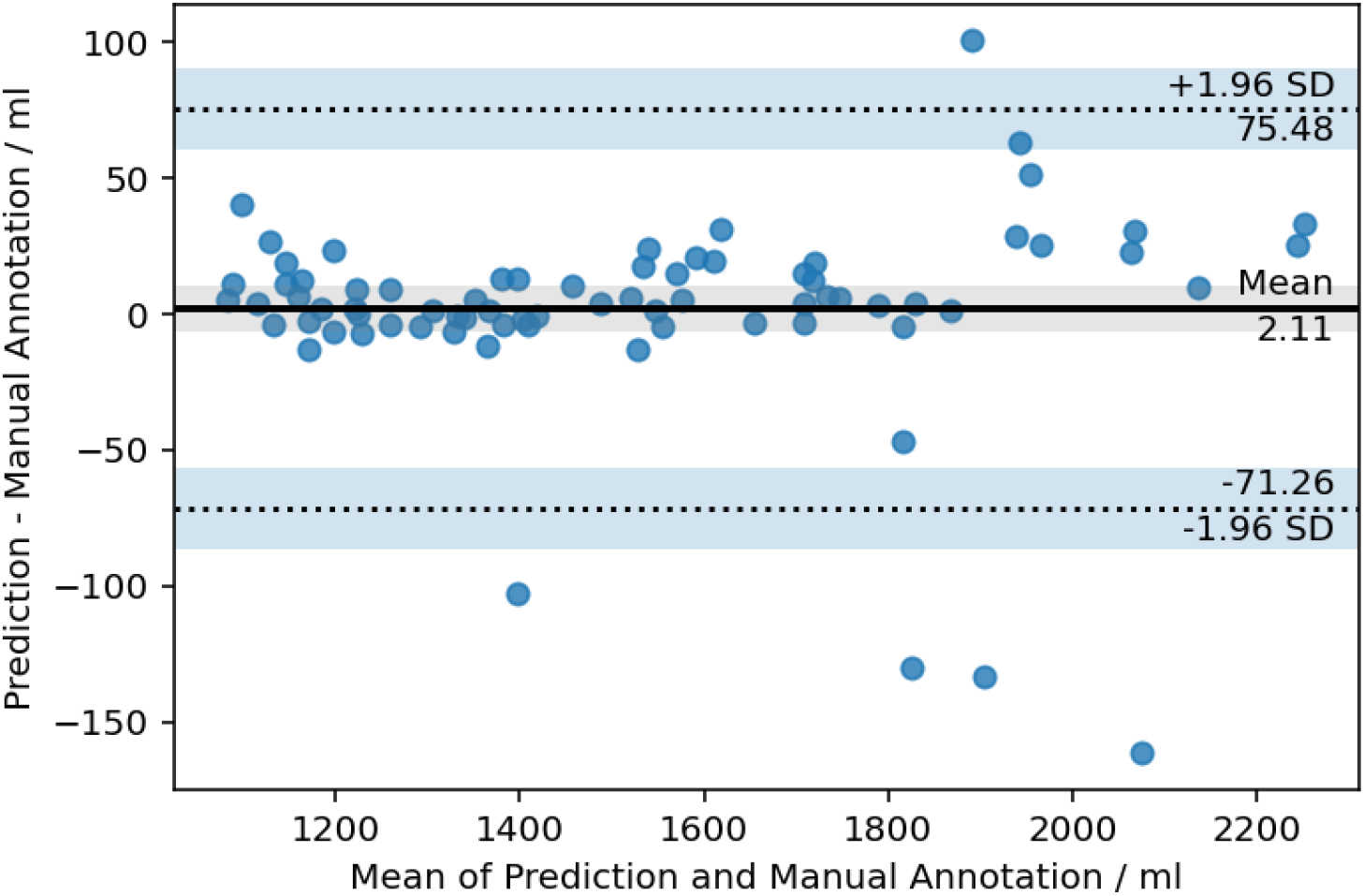
Bland-Altman plot for *Liver*

**Appendix 17:**
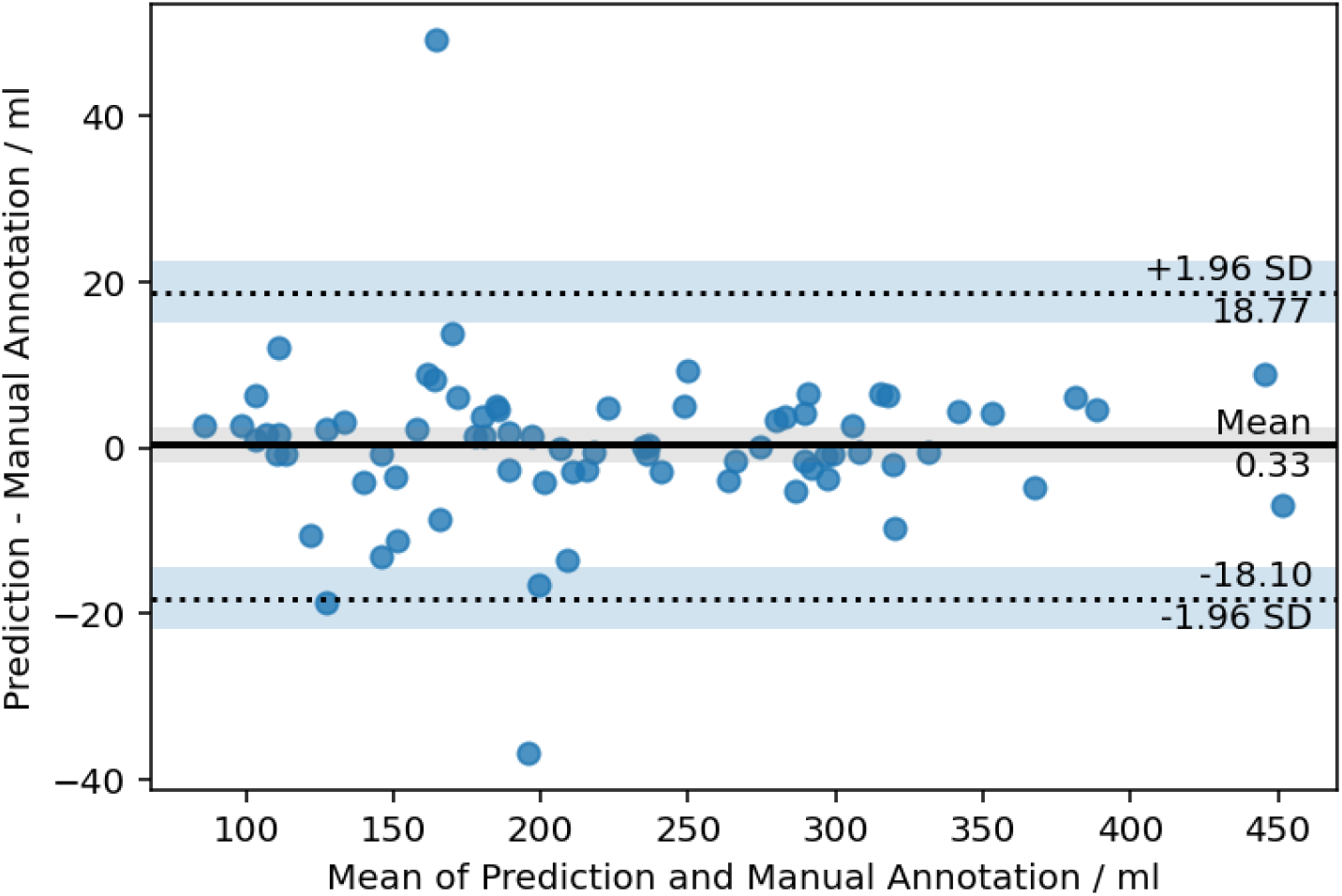
Bland-Altman plot for *Spleen*

**Appendix 18:**
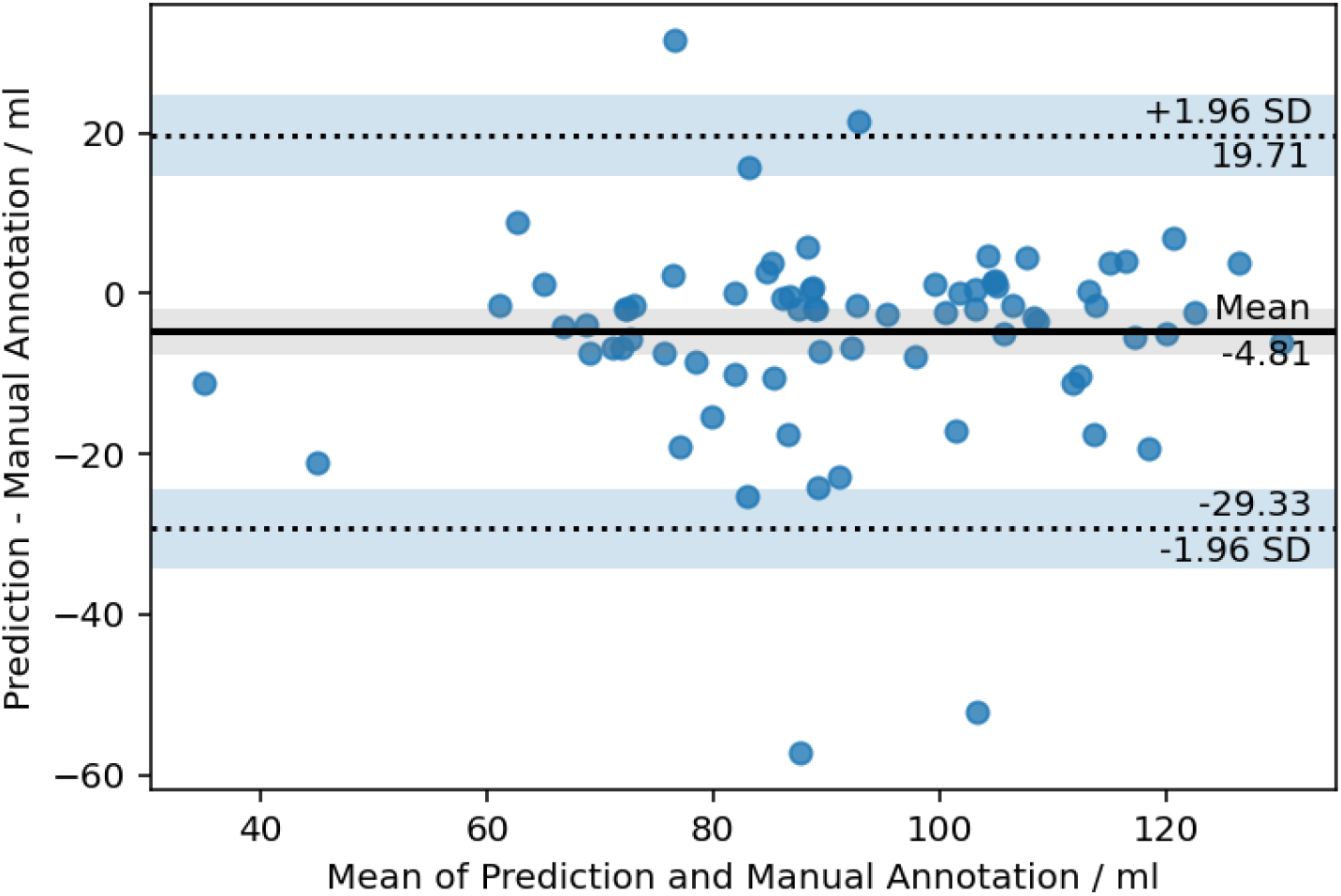
Bland-Altman plot for *Pancreas*

**Appendix 19:**
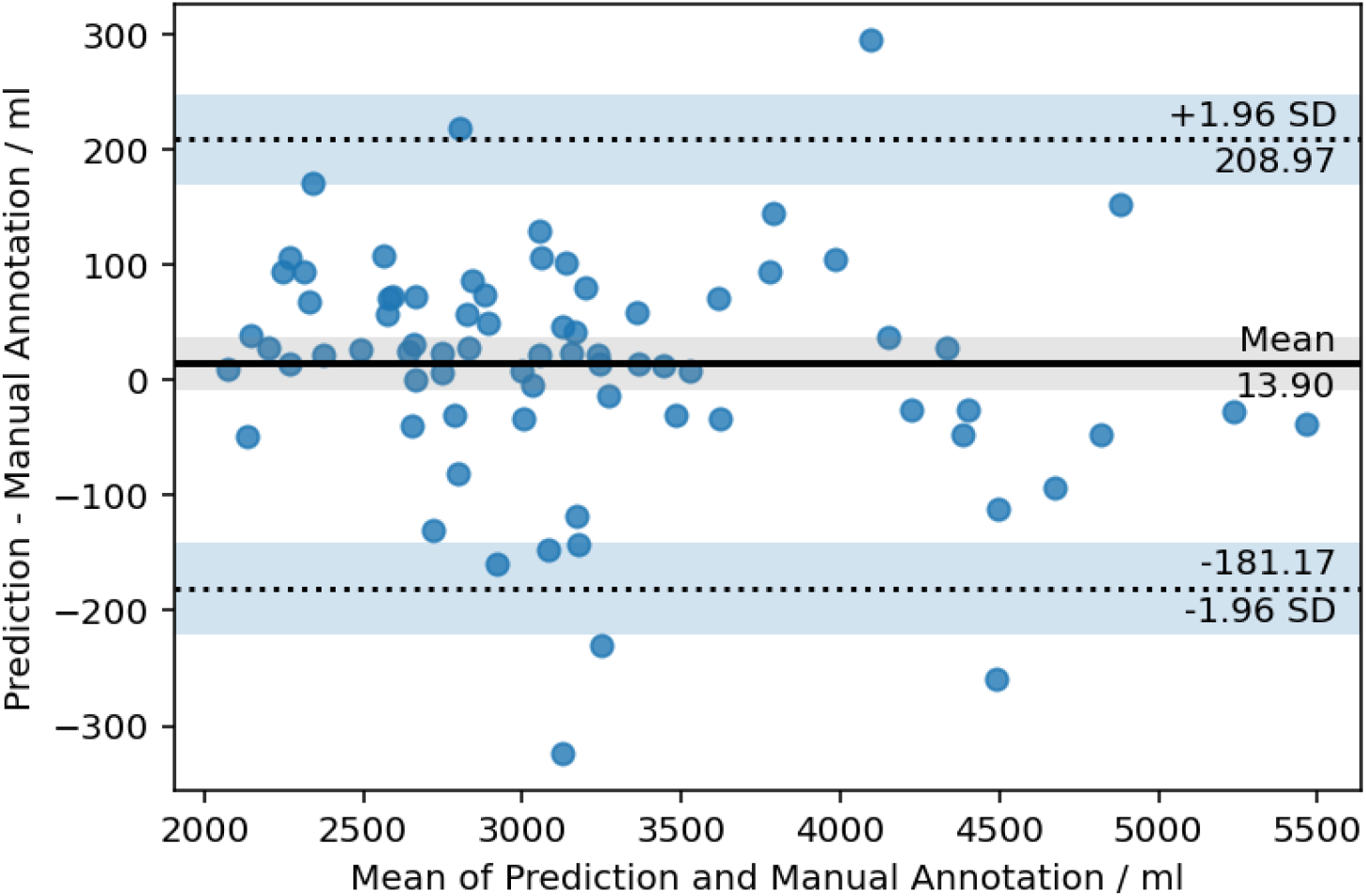
Bland-Altman plot for *Calves*

